# Estrogen Receptor Gene Expression Prediction from H&E Whole Slide Images

**DOI:** 10.1101/2024.04.05.24302951

**Authors:** Anvita A. Srinivas, Ronnachai Jaroensri, Ellery Wulczyn, James H. Wren, Elaine E. Thompson, Niels Olson, Fabien Beckers, Melissa Miao, Yun Liu, Po-Hsuan Cameron Chen, David F. Steiner

**Affiliations:** Google Health, Palo Alto, CA; Henry M. Jackson Foundation, Bethesda, MD; Defense Innovation Unit, Mountain View, CA; Naval Medical Center San Diego, San Diego, CA; Verily Life Sciences, South San Francisco, CA

## Abstract

Gene expression profiling (GEP) provides valuable information for the care of breast cancer patients. However, the test itself is expensive and can take a long time to process. In contrast, microscopic examination of hematoxylin and eosin (H&E) stained tissue is inexpensive, fast, and integrated into the standard of care. This work explores the possibility of predicting *ESR1* gene expression from H&E images, and its use in predicting clinical variables and patient outcomes. We utilized a weakly supervised method to train a deep learning model to predict *ESR1* expression from whole slide images, and achieved 0.57 [95% CI: 0.46, 0.67] Pearson’s correlation with the ground truth value. Our *ESR1* expression prediction achieved an AUROC of 0.81 [0.74, 0.87] in predicting clinical ER status obtained using an immunohistochemistry staining technique, and a c-index of 0.59 [0.52, 0.65] in predicting progression-free interval for the patients in our cohort. This work further demonstrates the potential to infer gene expression from H&E stained images in a manner that shows meaningful associations with clinical variables. Because obtaining H&E stained images is substantially easier and faster than genetic testing, the capability to derive molecular genetic information from these images may increase access to this type of information for patient risk stratification and provide research insights into molecular-morphological associations.

## 1 Introduction

Breast cancer is the second leading cause of cancer deaths among American women, with roughly 40,000 deaths each year [7]. The treatment of breast cancer is complex, with protocols that differ based on the many subtypes. These subtypes are traditionally determined by examining the tumor under a microscope. However, molecular testing, such as gene expression profiling (GEP) has also become an important source of information to inform treatment in some cases. For example, OncotypeDx, a GEP test, can identify patients with low risk cancer to help them make the decision to forgo chemotherapy and thus avoid the associated toxicities and costs of such treatment [10].

Despite its benefit, patients face challenges to access this type of molecular testing– the test costs several thousands of dollars, requires coordination of tissue shipping, and can take several weeks to return results. Furthermore, genetic testing can require a substantial amount of tissue, which may be an issue of particular importance for small tumors or low tumor content specimens. In this work, we explore predicting gene expression directly from images of hematoxylin and eosin (H&E) stained tissue. Since H&E staining is a fast and inexpensive component of the standard tissue preparation in pathology, this approach requires no additional tissue usage. Our focus is on expression of *ESR1*, the gene encoding estrogen receptor (ER) which is a key marker of breast cancer subtype and an informative component of the OncotypeDX test. We predict the expression of this gene using a weakly supervised method [3]. Going beyond previous work that demonstrated the feasibility of image based prediction of gene expression [9], we evaluate the use of the resulting *ESR1* gene expression prediction model to further predict clinical ER status and patient outcome.

## 2 Method

### 2.1 Dataset

We utilized two de-identified datasets in this work: 929 cases from the publicly available The Cancer Genome Atlas (TCGA) generated by the TCGA Research Network (https://www.cancer.gov/tcga), and 432 cases from a tertiary teaching hospital (TTH). Both datasets contained whole slide images (WSI) of H&E-stained pathology slides, ER status based on original clinical review of immunohistochemistry (IHC), and survival data [2, 5]; TCGA also contained gene expression data (RNASeqV2 with data provided as normalized counts using the RSEM method; log2 transformed for analysis; more detail is given in [4]). TCGA cases were split based on tissue source sites into 551, 199, and 179 cases for train, validation, and test respectively. TTH cases were used exclusively as a test set for evaluation of ER status and clinical outcome prediction (gene expression data not available for this dataset). The study was reviewed by the Advarra institutional review board (Columbia, Maryland) and deemed exempt from informed consent as all data were retrospective and de-identified.

### 2.2 Approach

The WSIs of breast cancer tissue can be 10^8^ or more pixels each, making it challenging to pass an entire image through a regression model on a single machine. We follow the Multiple Instance Learning (MIL) approach [3], which uses gated attention to aggregate patch-level image embeddings into a single prediction. Here, each image is assumed to be a bag of patches, with a single image-level floating-valued label representing the gene expression.

To obtain patch-level embeddings, we first sample 16,843 patches of size 224 *×* 224 at 10*×* magnification (1 *µ*m per pixel) from each image randomly without replacement. The patches are then passed through a pre-trained CNN that generates embedding of size 2,048. The CNN was pre-trained using contrastive learning on pathology images [1].

The patch embeddings are then fed through the MIL model to generate the final prediction. The MIL model consists of a fully connected layer and an attention layer of size 8 for the gated attention mechanism. This layer computes the weighted combination of all the patch embeddings of an image. The output of the model is a single unbounded floating number representing the predicted gene expression. The model was trained to minimize mean squared error using the Adam optimizer [6]. We tuned all hyperparameters based on the performance on the validation set.

## 3 Results

Table 1 shows our model performance in predicting gene expression based on root mean square error (RMSE), mean absolute error (MAE), and Pearson correlation with the ground truth values. As a baseline, we compute the dataset variance (i.e., “predicting” using the mean value). Our model achieved a RMSE, MAE, and correlation of 2.90 [95% CI: 2.57, 3.23], 2.20 [1.91, 2.49], 0.57 [0.46, 0.67], respectively.

**Table 1:**
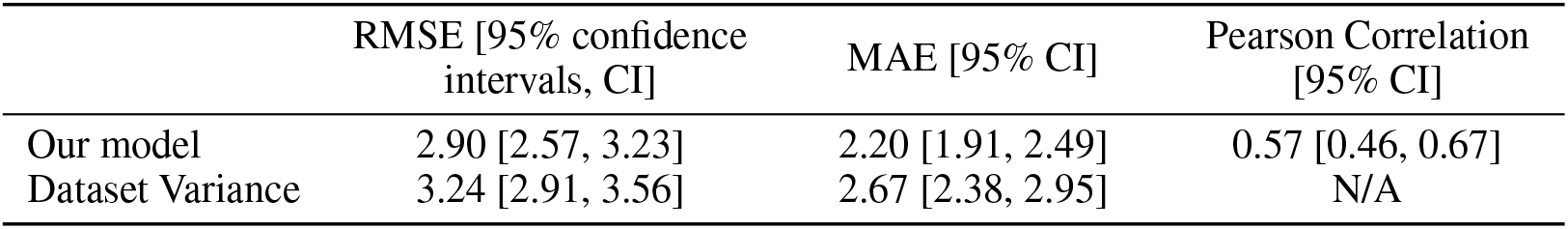
RMSE and MAE values for the MIL model on the TCGA test set, compared to a baseline of predicting the dataset mean.

**Table 2:** Balanced accuracy and AUROC for this work on the TCGA test set. We compare the results to a fully supervised method [2]. To get an upper bound for the performance we can expect from a *ESR1* regression model for the task of ER status prediction, we compute the metrics using the ground truth *ESR1* expression value.

| Metric | This work (using *ESR1* expression prediction to classify ER status) [95% CI] | Fully-supervised method for direct ER status classification [2] [95% CI] | Using ground truth *ESR1* expression to classify ER status (upper bound) [95% CI] |
| --- | --- | --- | --- |
| Balanced Accuracy | 0.78 [0.70, 0.85] | 0.76 [0.65, 0.86] | 0.94 [0.89, 0.97] |
| AUROC | 0.81 [0.74, 0.87] | 0.85 [0.77, 0.92] | 0.95 [0.91, 0.98] |

Next, we evaluated whether the model can meaningfully predict important clinical variables. ER status evaluation by immunohistochemistry (IHC) is part of the routine clinical workup for breast cancer. A pathologist examining the tumor using immunohistochemistry (IHC) stains determines the tumor to have positive or negative ER expression based on visual interpretation of staining intensity and distribution. Since *ESR1* gene expression is associated with ER (protein) expression, we expect *ESR1* value to be predictive of the ER status. We first evaluate how well our model can predict the ER status. This is done by thresholding the continuous predictions from our model to obtain either a ‘positive’ or a ‘negative’ ER status prediction. We use a threshold value of 10 based on the validation data, and find that our weakly supervised method based on regressing against *ESR1* is comparable to a fully-supervised method directly predicting ER status [2]. This is particularly interesting because the method proposed in [2] requires strong supervision based on extensive annotations, and a 2-stage modeling approach, whereas our approach is weakly supervised and therefore does not require expert labeling of *ESR1*/ER.

As patients with ER-positive tumors generally have a better prognosis as compared to those with ER-negative tumors, we also evaluated if *ESR1* expression is predictive of patient outcomes. We consider breast cancer progression (as indicated by recurrence and deaths) as the endpoint, and we measure the concordance index (C-index) using the *ESR1* prediction output directly as the risk scores (Table 3). Our model achieved a c-index of 0.59 [95% CI: 0.52, 0.65]. For comparison, the c-index using the clinical ER status was 0.61 [0.54, 0.66] for this dataset. The pathologist-provided tumor grade, another established pathologic factor associated with prognosis [8], had a c-index of 0.64 [0.58, 0.69]. Additionally, as Figure 1 shows, risk stratification using the binarized *ESR1* expression predictions could identify high and low risk patients with different outcomes (logrank test, p=0.018).

**Table 3:** Survival analysis on the TTH dataset. We compared the performance to those provided by pathologist-provided ER status, and pathologist-provided tumor grade.

|  | This work [95% CI] | IHC-based ER status [95% CI] | Pathologist-provided tumor grade [95% CI] |
| --- | --- | --- | --- |
| C-index | 0.59 [0.52, 0.65] | 0.61 [0.54, 0.66] | 0.64 [0.58, 0.69] |

**Figure 1:**
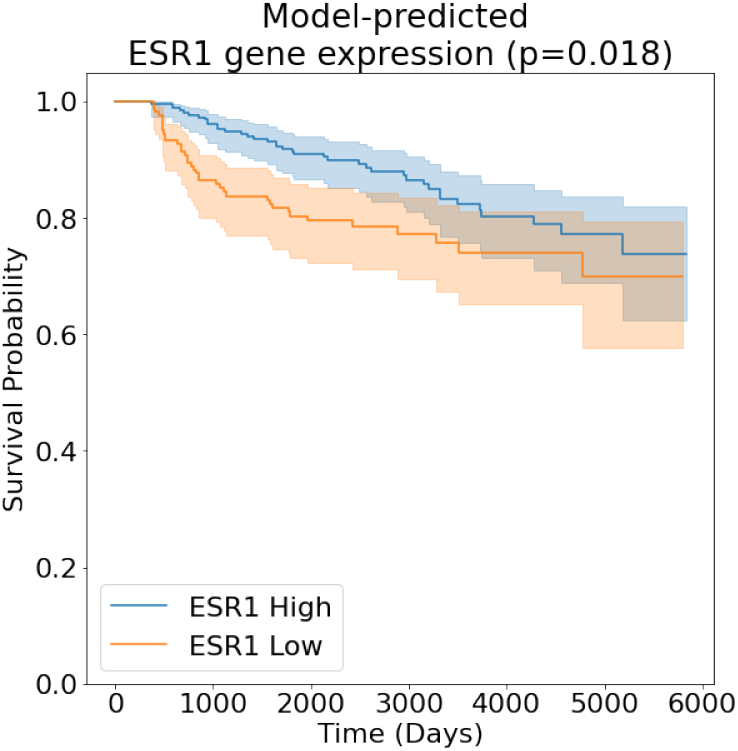
Kaplan-Meier curves for patients from the TTH dataset predicted to have tumors with high (blue) vs low (orange) *ESR1* expression. High and ^3^low classification are based on the threshold of 10 found on the TCGA validation set. Shaded areas represent 95% CIs; p-value is computed using the logrank test.

## 4 Discussion

Compared to genetic testing and gene expression profiling in particular, obtaining H&E stained images is substantially easier and faster. The capability to derive genetic information from these images may therefore increase access to genomic information, enabling clinicians to make more informed decisions about each patient’s treatment or providing insights about mechanisms of disease. This work is only a first step however, and further research on this topic will be needed to help this capability materialize. Limitations of this study include the fact that treatment data is not available for these datasets and treatment both influences outcomes and is in part determined by the ER status itself. A larger dataset, potentially with controlled treatment arms, is thus needed to validate the survival prediction power of the model. Additionally, the H&E images from TCGA are not necessarily immediately adjacent structurally to the tissue used for the molecular testing. Thus, heterogeneity in the tumor or the tumor content across slides from the same tumor may create noise between label and image regarding the values used for gene expression. Lastly, *ESR1* is only one gene out of thousands expressed in breast cancer. Since several studies [10, 11] demonstrate the power of multiple genes to predict patient response, additional genes should be included and studied to create a more robust prognostic panel.

## Data Availability

All data produced in the present study are available upon reasonable request to the authors

https://www.cancer.gov/ccg/research/genome-sequencing/tcga

